# High prevalence of long-term psychophysical olfactory dysfunction in patients with COVID-19

**DOI:** 10.1101/2021.01.07.21249406

**Authors:** Paolo Boscolo-Rizzo, Anna Menegaldo, Cristoforo Fabbris, Giacomo Spinato, Daniele Borsetto, Luigi Angelo Vaira, Leonardo Calvanese, Andrea Pettorelli, Massimo Sonego, Daniele Frezza, Andy Bertolin, Walter Cestaro, Roberto Rigoli, Giancarlo Tirelli, Maria Cristina Da Mosto, Anna Menini, Jerry Polesel, Claire Hopkins

## Abstract

This study prospectively assessed the long-term prevalence of self-reported and psychophysically measured olfactory dysfunction in subjects with mild-to-moderate COVID-19. Self-reported smell or taste impairment was prospectively evaluated by SNOT-22 at diagnosis, 4-week, 8-week, and 6-month. At 6 months from the diagnosis, psychophysical evaluation of olfactory function was also performed using the 34-item culturally adapted University of Pennsylvania Smell Identification Test (CA-UPSIT). 145 completed both the 6-month subjective and psychophysical olfactory evaluation. According to CA-UPSIT, 87 subjects (60.0%) exhibited some smell dysfunction, with 54 (37.2) being mildly microsmic, 16 (11.0%) moderately microsmic, 7 (4.8%) severely microsmic, and 10 patients (6.9%) being anosmic. At the time CA-UPSIT was administered, a weak correlation was observed between the self-reported alteration of sense of smell or taste and olfactory test scores (Spearman’s r=-0.26). Among 112 patients who self-reported normal sense of smell at last follow-up, CA-UPSIT revealed normal smell in 46 (41.1%), mild microsmia in 46 (41.1%), moderate microsmia in 11 (9.8%), severe microsmia in 3 (2.3%), and anosmia in 6 (5.4%) patients; however, of those patients self-reporting normal smell but who were found to have hypofunction on testing, 62 out of 66 had self-reported reduction in sense of smell or taste at an earlier time point. Despite most patients report a subjectively normal sense of smell, we observed a high percentage of persistent smell dysfunction at 6 months from the diagnosis of SARS-CoV-2 infection, with 11.7% of patients being anosmic or severely microsmic. These data highlight a significant long-term rate of smell alteration in patients with previous SARS-CoV-2 infection.

## Introduction

In patients with mildly symptomatic coronavirus disease 2019 (COVID-19), sudden changes in the sense of smell or taste are reported by around two-thirds of subjects (Spinato et al., 2020) being an early marker of COVID-19 and representing the most common long-lasting symptom of severe acute respiratory syndrome coronavirus 2 (SARS-CoV-2) infection (Boscolo-Rizzo et al., 2020c).

Considering the high spreading of SARS-CoV-2 infection worldwide, COVID-19 is expected to significantly contribute to the overall burden of anosmia in next future. Thus, it is imperative to systematically monitor the olfactory and gustative function in this population (Vaira et al., 2020a).

To date, only a few authors have investigated the recovery of the olfactory function in COVID-19 patients with psychophysical tests (Lechien et al., 2020; Vaira et al., 2020b). However, none of these studies exceeded the 2-month follow-up and therefore further improvement may occur. To the best of our knowledge, only two studies, based on data obtained from the patient’s interview, have so far investigated the recovery of olfactory dysfunction 6 months after onset (Hopkins et al., 2020; Klein et al., 2020).

We have previously reported the prevalence of altered smell or taste in a cohort of mildly symptomatic home-isolated patients with confirmed SARS-CoV-2 infection (Spinato et al., 2020) as well as the evolution of these symptoms at 4 (Boscolo-Rizzo et al., 2020a) and 8 weeks (Boscolo-Rizzo et al., 2020c). This study aims to assess the 6-month prevalence of self-reported and psychophysically measured olfactory dysfunction using a modified Italian version of the University of Pennsylvania Smell Identification Test (UPSIT) in that cohort of patients.

## Materials and Methods

### Subjects

We conducted a prospective study on adult patients consecutively assessed at Treviso Regional Hospital who tested positive for SARS-CoV-2 RNA by polymerase chain reaction (PCR) on nasopharyngeal and throat swabs performed according to World Health Organization recommendation (World Health Organization 2020). All patients were initially home-isolated with mild-to-moderate symptoms. Patients were considered mildly symptomatic if they had less severe clinical symptoms with no evidence of pneumonia, not requiring hospitalization, and therefore considered suitable for being treated at home. Patients with a history of previous craniofacial trauma, surgery or radiotherapy in the oral and sinonasal area, and those reporting a pre-existing olfactory dysfunction were excluded from the study.

The study was conducted with the approval of the institutional ethical review boards of Treviso and Belluno provinces and informed consent was obtained verbally and in written form.

### Questionnaires

After collecting clinical data through a survey administered by telephone interview at the time of diagnosis, the same patients were re-contacted after 4 and 8 weeks. During the subsequent interviews the same questions were re-administered. In details, symptoms were assessed through *ad hoc* questions and structured questionnaires, including the ARTIQ (Acute Respiratory Tract Infection Questionnaire) and the SNOT-22, item “Sense of smell or taste”, as previously reported (Spinato et al., 2020). Briefly, the SNOT-22 grades symptom severity as none (0), very mild (1), mild or slight (2), moderate (3), severe (4), or as bad as it can be (5).

### Olfactory Testing

Finally, at 6-months from diagnosis, the Italian version of the UPSIT (Sensonics, Haddon Heights, NJ, USA) was used to assess the ability to identify odorants (Doty et al., 1984b) along with the administration of the above described questionnaires. The UPSIT is a well validated and reliable scratch-and-sniff test (test-retest r=0.94) based on the forced-choice among four alternative odorants. According to manufacturer instructions, each patient was asked to scratch each of the 40 microencapsulated odorants present in the booklets with the tip of a pencil, provided in the kit, and to select the name of the odorant closest to the one perceived. Each patient was assisted by an examiner to verify that the test was correctly performed. The number of correct answers was calculated according to the test manual. As the names of 6 odorants reported in the UPSIT test have previously been shown not to match the common perception of those odorants by the Italian population leading to misidentification > 20% in a population of normal subjects, a reduced and culturally adapted version of the Italian UPSIT (CA-UPSIT) including 34 odorants that had been identified correctly by more than 80% of normal subjects, was used to evaluate the sense of smell in our subjects according to Cenedese *et al*. (Cenedese et al., 2015) and Parola *et al*. (Parola and Liberini, 1999). The following scores on a scale of 0-34 have been used: probable malingering, 00-04; total anosmia, 05-15; severe microsmia, 16-19; moderate microsmia, 20-23; mild microsmia, 24-27; normosmia, 28-34.

Patients who refused to perform UPSIT test were asked to complete the 6-months questionnaires by telephone interview.

### Statistical analysis

Symptom prevalence was expressed as percentage of total patients, and 95% confidence interval (CI) were calculated using Clopper-Pearson method and differences in prevalence were evaluated through Fisher’s exact test. Correlation between CA-UPSIT score and SNOT-22 was evaluated through Spearman’s correlation coefficient (r) and concordance between SNOT-22 at different follow-ups was evaluated through weighed Cohen’s kappa coefficient. The trend in mean of CA-UPSIT score across SNOT-22 categories were evaluated through ANOVA with constrains for trend. Statistical analyses were performed using R 3.6.

## Results

### Evolution of self-reported altered sense of smell or taste

Of 202 mildly symptomatic SARS-CoV-2 RNA positive adults consecutively assessed at Treviso Regional Hospital and completing the survey at the baseline, 183 subjects (90.6%) answered to both the 4-week, 8-week, and 6-month follow-up interview.

Symptom’s evolution from baseline to 6-month follow-up are reported in Table 1. Fatigue (19.7%), breathing problems (16.4%), and altered sense of smell or taste (18.0%) were by far the predominant long-lasting symptoms.

**Table 1.**
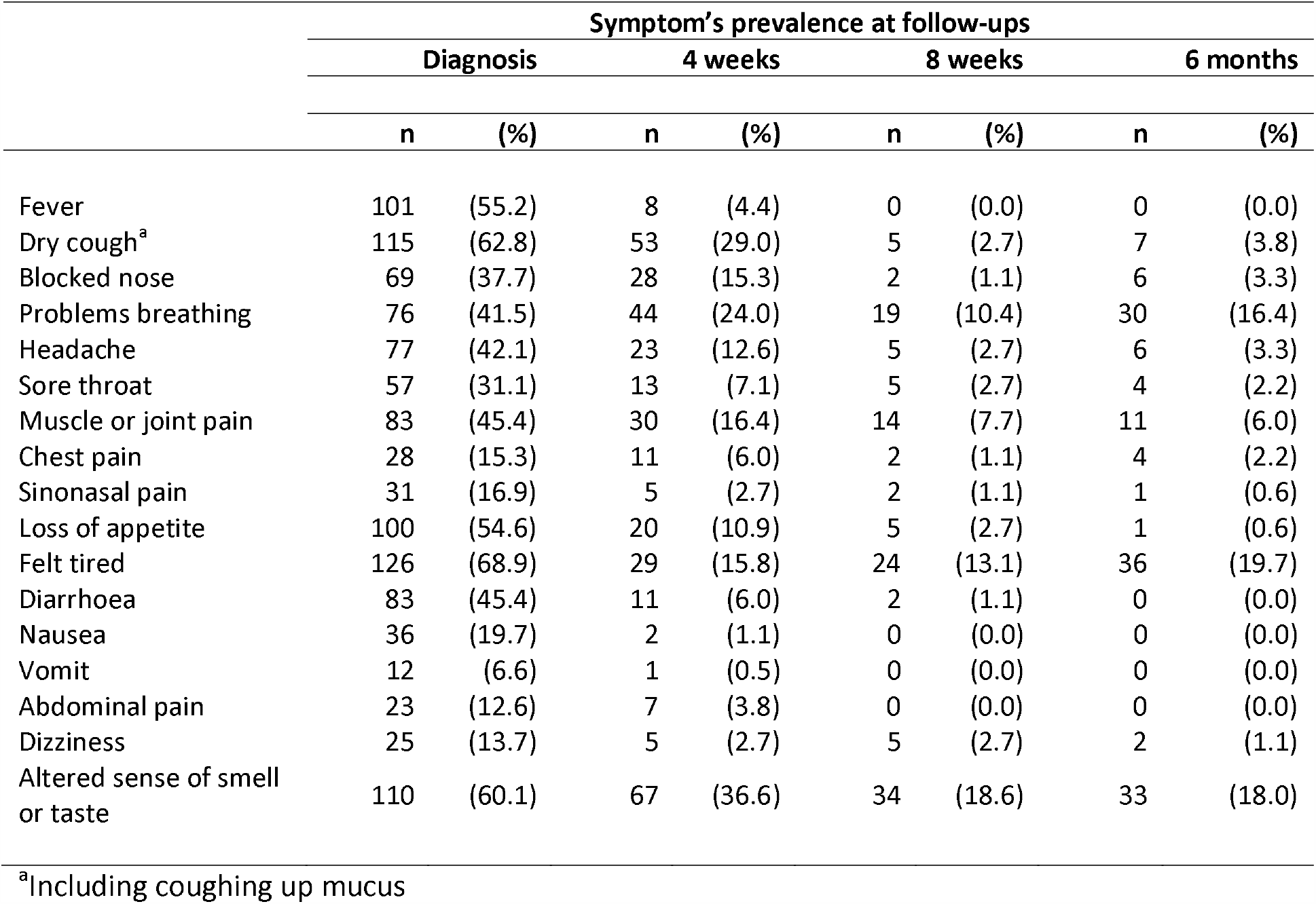
Symptom’s evolution from diagnosis to 6-month follow-up in 183 patients with complete follow-up.

The evolution of self-reported altered sense of smell or taste according to SNOT-22 score is shown in Figure 1 and in Appendix 1. Compared to the baseline, the prevalence of subjects complaining an altered sense of smell or taste dropped from 60.1% to 18.0% (Figure 1). Particularly at 6-months, among the 110 patients complaining a sudden onset of altered sense of smell or taste at baseline (Appendix 1), 85 (77.3%) self-reported a complete resolution of these symptoms, 22 (20.0%) a partial improvement, and in 3 subjects (2.7%) the symptoms were unchanged or worse. Among the 73 patients who did not self-report an altered sense of smell or taste at baseline, 8 (11.0%) reported this symptom at 6-month follow-up. Minor symptomatic changes have been observed from the 8-week to the 6-month follow-up (Cohen’ kappa=0.511 – Appendix 1), with 18 (9.8%) patients reporting symptomatic improvement and 19 patients (10.4%) reporting a worsening in the intensity of chemosensory alteration.

**Figure 1.**
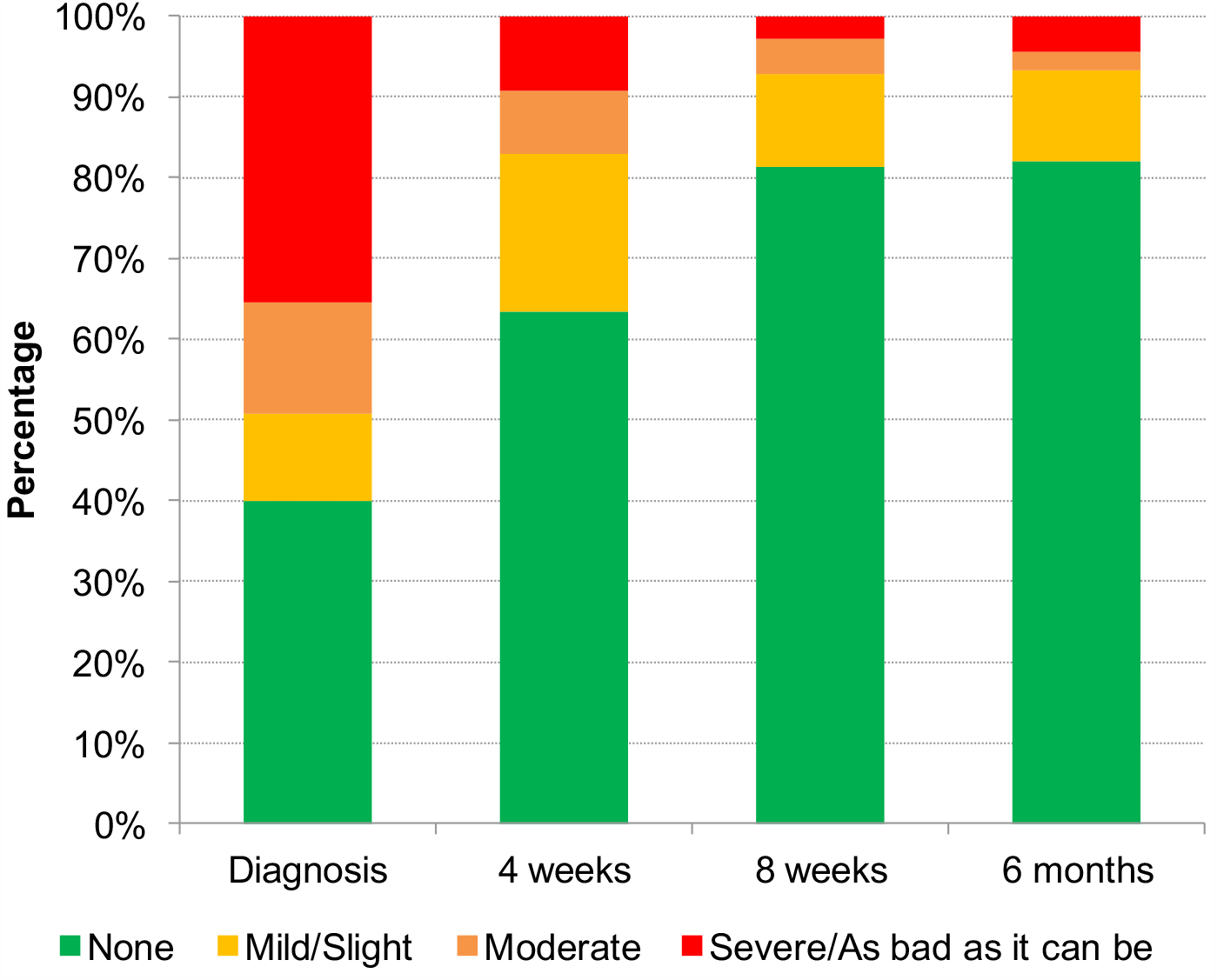
Evolution of alteration of sense of smell or taste (SNOT-22) in 183 patients with complete follow-up.

### Psychophysical olfactory evaluation using CA-UPSIT

Among 183 patients that completed all follow-up interviews, 145 (79.2%) underwent the psychophysical olfactory evaluation at 6-month through CA-UPSIT; of these, 80 (55.2%) were female and the median age was 55 years (range: 21-84 years) with 84.8% of patients aged ≤ 69 years. There were no significant socio-demographic differences and SNOT-22 score at baseline between patients completing the 6-month psychophysical evaluation and those who did not.

The mean CA-UPSIT score was 25.5 (SD=±5.4). According to the score, 87 subjects (60.0%) exhibited some smell dysfunction, with 54 (37.2) being mildly microsmic, 16 (11.0%) moderately microsmic, 7 (4.8%) severely microsmic, and 10 patients (6.9%) being anosmic (Table 2). Olfactory dysfunction was associated with older age but not with gender (Table 2). In patients who reported alteration of smell or taste either at 0, 4 or 8 weeks, the mean CA-UPSIT score was 25.3 (±5.8) and 26.1 (±4.3) in patients who have never reported smell loss (*P*=0.350).

**Table 2.**
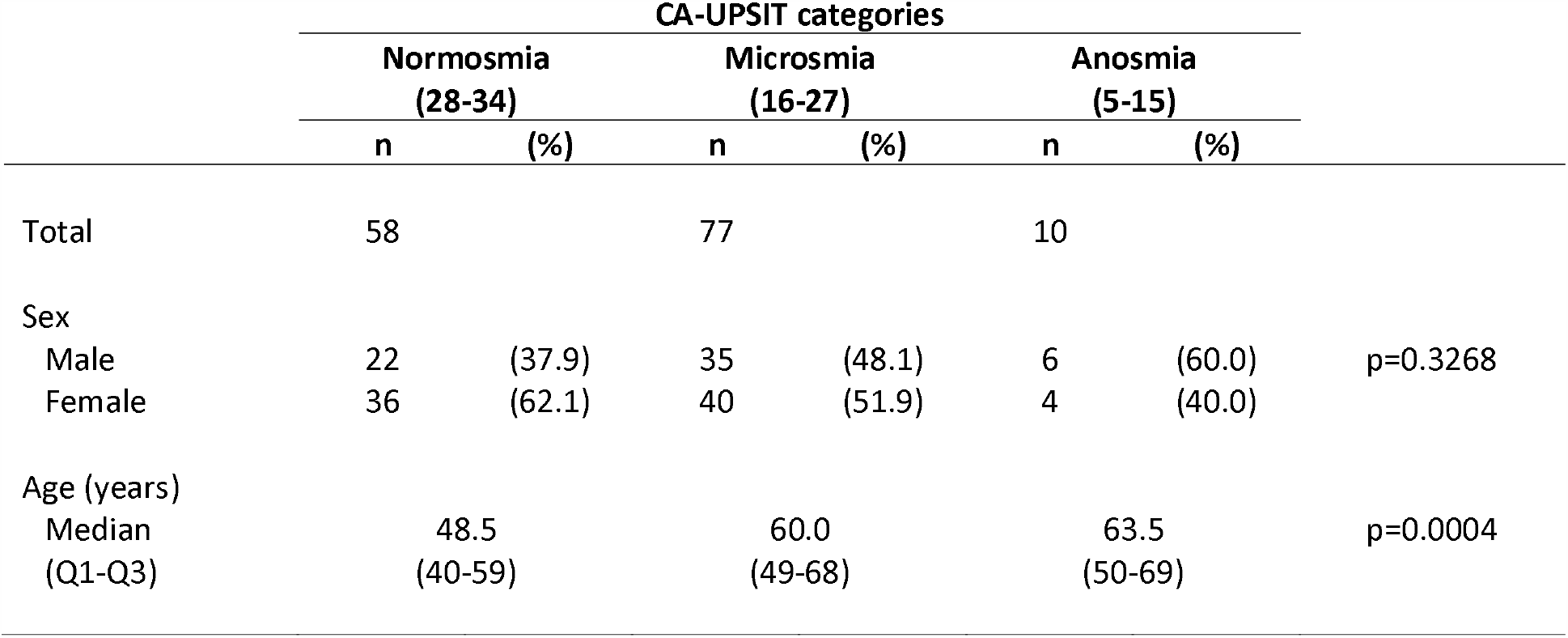
Six-month follow-up: alteration of sense of smell according to CA-UPSIT accortding to socio-demographic characteristics.

At the time CA-UPSIT was administered, a weak correlation was observed between the self-reported alteration of sense of smell or taste and olfactory test scores (Spearman’s r=-0.26). The mean CA-UPSIT score in patients with the score for self-reported loss of smell or taste ‘None’ was 26.3 ±4.8 vs 14.9 ±7.1 in patients self-reporting loss of smell as “Severe”/”As bad as it can be” (Figure 2; p<0.0001).

**Figure 2.**
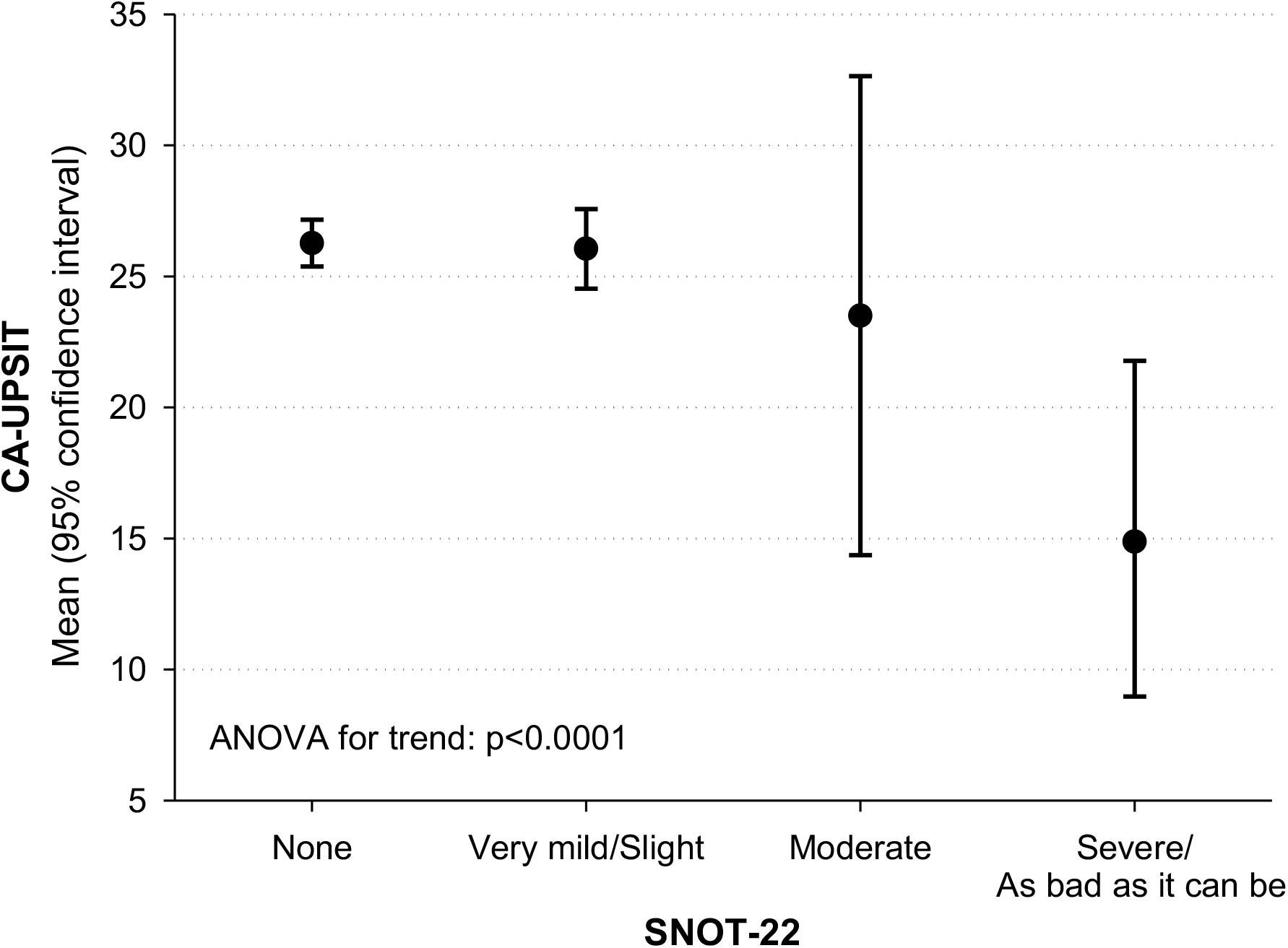
Six-month follow-up: mean CA-UPSIT and corresponding 95% confidence interval according to SNOT-22.

Among 145 patients undergoing CA-UPSIT evaluation, 112 self-reported normal sense of smell at 6-months follow-up; in these patients, CA-UPSIT revealed normal smell only in 46 (41.1%), while a mild microsmia was detected in 46 (41.1%), moderate microsmia in 11 (9.8%), severe microsmia in 3 (2.3%), and anosmia in 6 (5.4%) patients. CA-UPSIT revealed microsmia/anosmia in 66 patients who self-reported normal sense of smell or taste according to SNOT-22. However, of those patients self-reporting normal smell but who were found to have hypofunction on testing, 62 out of 66 (93.9%) had self-reported reduction in sense of smell or taste at an earlier time point.

Percentages of subjects who correctly identified each item of the CA-UPSIT according to impairment of sense of smell are reported in Figure 3. With the exception of “Pizza”, “Smoke”, and “Pine”, all other items showed significant differences in the discrimination rate when comparing the group of anosmic and normosmic patients with “Peach”, “Watermelon”, “Cinnamon”, “Menthol”, and “Coconut” being the odors with the lower rate of discrimination in the group of subjects classified as anosmic according to the overall CA-UPSIT score.

**Figure 3.**
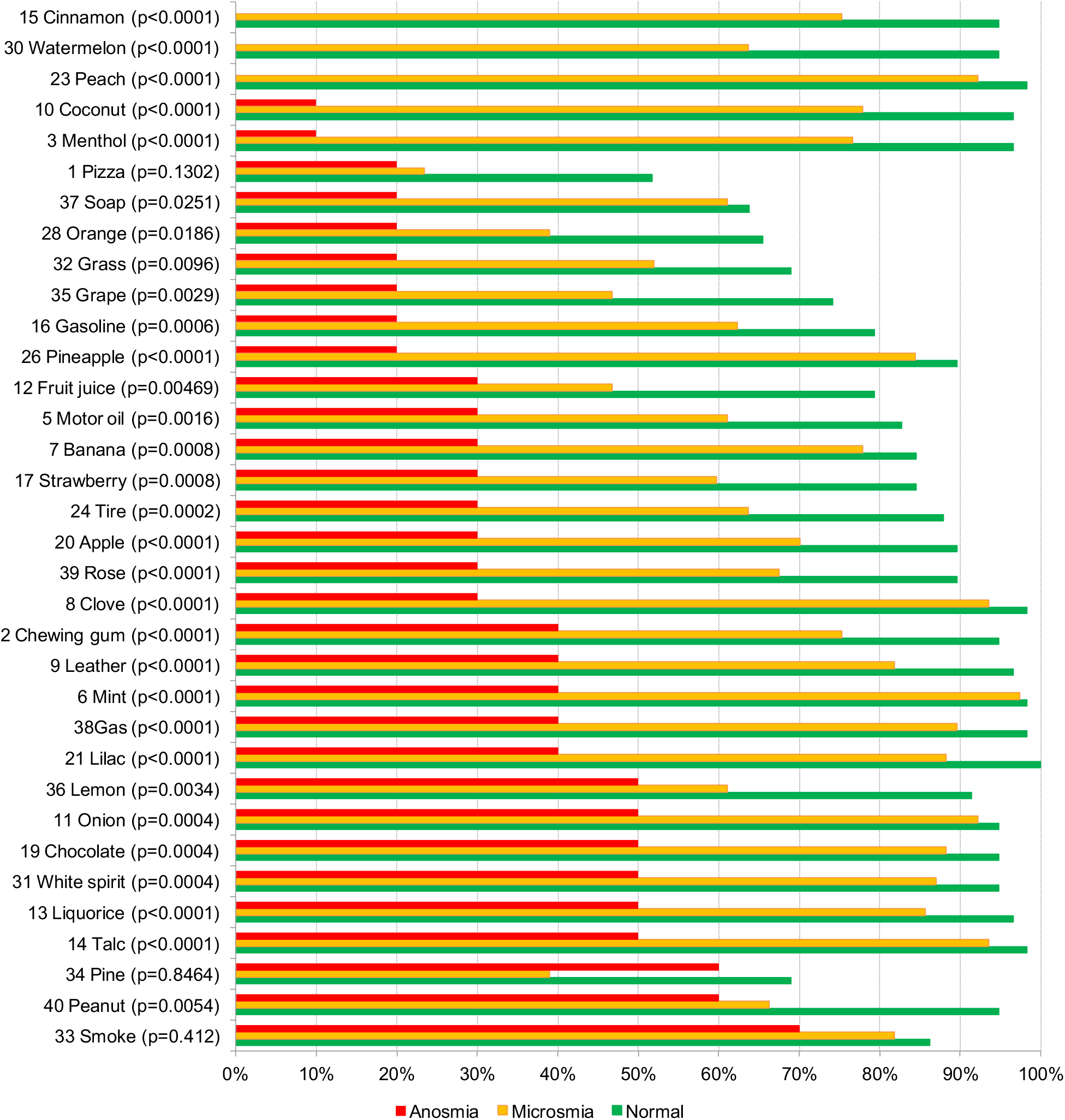
Identification of 34 odorants by level of impairment of sense of smell according to CA-UPSIT.

## Discussion

60.1% of our patient cohort reported loss of sense of smell or taste at the initial assessment shortly after COVID-19 diagnosis. Based on the subjective evaluation, 77.3% of these reported a complete resolution of these symptoms 6 months after the onset. We found that 6 months after SARS-CoV-2 infection, 60.0% of our patient cohort were found to have olfactory dysfunction with psychophysical evaluation, while only 18.0% of patients of this group self-reported an altered sense of smell or taste at the same time. This latter percentage is in line with another study that observed a 6-month follow-up prevalence of self-reported chemosensory changes of 14% in subjects with RT-PCR confirmed SARS-CoV-2 infection (Klein et al., 2020).

The quantitative evaluation of olfactory function was performed using CA-UPSIT, a modified version of the Italian scratch-and-sniff UPSIT containing 34 (instead of 40) odorants that are well recognized by Italian subjects (Parola and Liberini, 1999; Cenedese et al., 2015). The present study identified that at 6-month after SARS-CoV-2 infection, the average number of correct responses was 25.5 with 40.0% of patients being normosmic. 60.0% of patients had some quantitative reduction in olfactory function with 11.7% subjects being severely microsmic or anosmic.

Published control data on healthy Italian subjects of age comprised between 21 and 57 years reported a mean CA-UPSIT score of 32 with 98% normosmic and 2% mild microsmic subjects (Cenedese et al., 2015). Take into account that smell identification ability declined markedly only after the seventh decade (Doty et al., 1984a) and that only 15.2% of patients in the present series aged older than 69 years, our data confirm the presence of a significant olfactory impairment at 6-month.

Although we have encouraged the self-reported evaluation of anosmia as it is widely available and has as a baseline parameter of comparison (that is, the subjective perception of smell preceding the onset of COVID-19 (Boscolo-Rizzo et al., 2020b), these observations suggest that a subjective evaluation of the olfactory function may be inadequate to evaluate olfactory recovery. Consistently, several authors have underlined that subjectivity of self-reporting may lead to underestimation of the prevalence of olfactory dysfunction (Mazzatenta et al., 2020; Moein et al., 2020; Vaira et al., 2020b; Yan et al., 2020). Even considering that the evaluation by psychophysical tests could overestimate the prevalence of COVID-19-related smell disorders as it can detect pre-existing smell alterations not perceived by the patient and not related to SARS-CoV-2 infection, the rate of psychophysical long-term olfactory dysfunction observed in the present series is significantly higher than 15% of no self-reported psychophysical olfactory impairment described in population-based study (Murphy et al., 2002).

Interestingly, 93.9% of patients showing some degree of olfactory dysfunction at psychophysical evaluation but self-reporting a normal sense of smell or taste at 6-month, have complained of a subjective chemosensory alteration during the course of SARS-CoV-2 infection. As observed in another study (Otte et al., 2020), this suggests that patients, who previously had a significant chemosensory alteration and had only a partial recovery, tend to overestimate the extent of their recovery thus reporting a normal although psychophysically still impaired olfactory function.

Remarkably, according to CA-UPSIT scores, 11.7% of subjects showed a psychophysical significant alteration in the sense of smell consisting in anosmia or severe hyposmia. Thus, considering the high prevalence of COVID-19 in the worldwide population, a parallel high incidence of long-term morbidity with significant impact on the quality of life of patients is expected in the near future as well as a significant burden of disease on the health system. This should increase efforts to search for therapeutic strategies that should be guided basis on the pathogenesis of the disease.

Several possible mechanisms for loss of smell in the course of COVID-19 have been explored but it is still unclear how SARS-CoV-2 mediates smell loss. Briefly, the pathogenesis of the olfactory dysfunctions can be explained by two theories of direct viral cytopathy and systemic inflammatory cascade of events. According to recent studies, SARS-CoV-2 may affect directly the olfactory epithelium through perturbation of supporting cells which maintain the integrity of the olfactory sensory neurons and express ACE2 protein and cell surface protease TMPRSS2 which are both necessary for the virus entry into the target cells (Brann et al., 2020; Fodoulian et al., 2020). However, systematic studies on the histopathology of the olfactory epithelium in patients with COVID-19 are lacking (Deshmukh et al., 2020). A preliminary report suggests a possible role of the inflammation in the pathogenesis of the olfactory loss thus providing rationale for use of corticosteroids (Vaira et al., 2020c). Furthermore, olfactory training is the only disease-specific intervention with demonstrated efficacy for the treatment of post-infectious olfactory dysfunction and it is assumed that it acts through by increasing both the regenerative ability and the neuroplastic potential of olfactory neurons. Therefore, given the above observations, the absence of randomized controlled trials investigating specific treatment in COVID-19-related anosmia, and the relatively high safety profile associated with their use, topical corticosteroid and olfactory training may be considered in selected patient with post-COVID-19 smell disturbance (Levy, 2020). Based on the evolution of the self-reported alteration of sense of smell or taste in the present series, symptoms tend to stabilize after two months from the onset. This could suggest that any therapy should be started early after the nasopharyngeal swab results negative as the possible adverse effects of a corticosteroid treatment in patients with ongoing upper airway SARS-CoV-2 infection remain to be investigated.

There are a number of limitations to this study. First, a baseline or early psychophysical evaluation of the sense of smell was not done in the present series. However, as previously discussed, despite the possibility that psychophysical tests could detect a pre-existing olfactory dysfunction not perceived by the patient, this may only partially impact on the rate of psychophysical long-term smell alteration observed in the present series. Second, a culturally adapted version of the Italian UPSIT was used to psychophysically evaluate the sense of smell. Despite this is the most reliable and accurate olfactory test available (Doty et al., 1984), it is only an identification test not testing discrimination ability and olfactory threshold. Thus, UPSIT identification test may not capture all olfactory disturbances.

However, it has been observed that patients with post-infectious hyposmia performed relatively well in both threshold and discrimination but poorly in identification tests (Liu et al., 2020). Identification testing alone may therefore over-estimate the severity of loss and may in part explain the weak correlation between self-reported smell loss and the UPSIT scores reported herein. Third, a psychophysical evaluation of the sense of taste was not performed in the present study. Although the commonly reported loss of taste might largely depend on impairment of retronasal olfaction, recent subjective studies have shown that COVID-19 associated chemosensory impairment is not limited to smell but also affects taste and chemesthesis (Gerkin et al., 2020; Parma et al., 2020). The psychophysical evaluation of the sense of taste as well as histopathological evaluation of taste buds and oral mucosa are highly desirable to clarify this aspect. Finally, these results apply to a subgroup of COVID-19 patients with mild-to-moderate disease. A long term psychophysical evaluation of the olfaction should be performed also in patients previously hospitalized for severe COVID-19 which seem to self-report a lower prevalence of smell or taste alteration at the onset of the disease (Borsetto et al., 2020). Finally, the abbreviated 34 item CA-SIT test has not been subjected to the same level of validation as the original UPSIT tests and this may reduce the reliability of scores.

## Conclusion

In conclusion, the results of the present study show a very high prevalence of long-term psychophysical olfactory dysfunction after SARS-CoV-2 infection. The large discrepancy between psychophysical and self-reported altered sense of smell highlights the importance to use psychophysical tests in the long-term follow-up of these patients to capture the burden of permanent olfactory dysfunction. Finally, there is a clear need for more research on treatment strategies for this group of patients.

## Data Availability

All relevant data are within the paper and its Supporting Information files.

## Availability of data and material

Additional informed consent was obtained from all individual participants for whom identifying information is included in this article

## Code availability

n/a

## Ethics approval

The study was conducted with the approval of the ethic committee for clinical experimentation of Treviso and Belluno provinces (ethic vote: 780/CE).

