## Appendix for "High prevalence of long-term psychophysical olfactory dysfunction in patients with COVID-19"

**Appendix 1**. Evolution of loss of sense of smell or taste from baseline to 6-month follow-up.

| **SNOT-22** | **SNOT-22 at 6-month follow-up** | | | | | | **Total** |
| --- | --- | --- | --- | --- | --- | --- | --- |
|  | **None** | **Very Mild** | **Slight** | **Moderate** | **Severe** | **As bad as**  **it can be** |  |
| Total | 150 | 14 | 7 | 4 | 5 | 3 | 183 |
| **Diagnosis** |  |  |  |  |  |  |  |
| None | 65 | 2 | 1 | 2 | 2 | 1 | 73 |
| Very mild | 5 | 0 | 0 | 0 | 0 | 0 | 5 |
| Slight | 13 | 2 | 0 | 0 | 0 | 0 | 15 |
| Moderate | 19 | 4 | 1 | 0 | 1 | 0 | 25 |
| Severe | 21 | 1 | 2 | 0 | 0 | 0 | 24 |
| As bad as it can be | 27 | 5 | 3 | 2 | 2 | 2 | 41 |
|  | Weighted Cohen’s kappa: 0.054 (-0.012-0.119) | | | | | |  |
| **8-week follow-up** |  |  |  |  |  |  |  |
| None | 139 | 5 | 2 | 2 | 0 | 1 | 149 |
| Very mild | 7 | 4 | 3 | 1 | 0 | 0 | 15 |
| Slight | 2 | 1 | 1 | 0 | 2 | 0 | 6 |
| Moderate | 2 | 2 | 1 | 0 | 1 | 2 | 8 |
| Severe | 0 | 2 | 0 | 1 | 2 | 0 | 5 |
| As bad as it can be | 0 | 0 | 0 | 0 | 0 | 0 | 0 |
|  | Weighted Cohen’s kappa: 0.511 (0.378-0.643) | | | | | |  |
